# Current tobacco use and associated factors among school-going adolescents in the Republic of Congo: Secondary analysis of the 2019 Global Youth Tobacco Survey and development of a simple identification score

**DOI:** 10.64898/2026.10.20.26366060

**Authors:** Esthel Lee Presley Bemba, Malonga Miafouna Axhel Ronan, Mbou Essie Darius Eryx, Okemba Okombi Franck Hardain, Batchi-Bouyou Armel

## Abstract

Data on adolescent tobacco use in the Republic of Congo are scarce, despite a 2012 tobacco control law and 2018 decrees banning smoking in public places and tobacco advertising, promotion and sponsorship. This study aimed to estimate current tobacco product use among Congolese middle school students, identify associated factors, and develop a simple score to identify current use. Secondary analysis of the cross-sectional 2019 Global Youth Tobacco Survey (GYTS). Current use was defined as self-reported use, in the past 30 days, of cigarettes (CR7), other smoked tobacco products (CR10) or smokeless tobacco (CR14). The estimates and regression models accounted for the sampling weights, 34 strata and 70 primary sampling units. A point-based score was developed from eight questionnaire-based factors and evaluated via cross-validation, which was grouped by primary sampling unit. Among the 6,297 students with a usable tobacco use status, 842 reported current use; the weighted prevalence was 13.5% (95% CI: 11.1–16.3%). The full model included 5,251 students, 597 of whom were current users. Being offered free tobacco by a tobacco company representative (aOR = 3.05), exposure to smoke at home (aOR = 2.71), seeing teachers smoke at school (aOR = 2.60) and exposure to smoke in enclosed public places (aOR = 2.02) were associated with current use. The score, ranging from 0 to 18 points, had a weighted AUROC of 0.771 in the development data and 0.765 in the grouped cross-validation data. Current tobacco use affects a substantial proportion of middle-school students. The associations with social and commercial exposures support the strengthening of prevention and the enforcement of existing legislation. The score is exploratory and requires external validation before any operational use.

## Introduction

Tobacco remains one of the leading preventable causes of death worldwide. According to the World Health Organization (WHO), it kills more than 7 million people each year, including approximately 1.6 million nonsmokers exposed to secondhand smoke. Approximately 80% of the world’s 1.3 billion tobacco users live in low- and middle-income countries [1]. Adolescence is the critical period of this epidemic: the vast majority of adult smokers started smoking before the age of 18, and nicotine dependence develops faster and more durably earlier [2]. Preventing tobacco initiation among young people is therefore the most effective lever for reducing, in the long term, tobacco-related morbidity and mortality, particularly respiratory and cardiovascular disease.

Globally, approximately one in eight adolescents aged 13–15 years uses a tobacco product [3]. While consumption is declining in high-income countries, sub-Saharan Africa is emerging as a region at risk. Its population is the youngest in the world, and the tobacco industry regards it as an expansion market. Tobacco control policies are still unevenly enforced. A pooled analysis of GYTSs from 22 African countries estimated that the prevalence of current tobacco use among school-going adolescents was 19.1% [4]. This work also highlighted the growing role of products other than cigarettes, such as smokeless tobacco, waterpipes (shisha) and other smoked tobacco, which often escape prevention messages focused on cigarettes [4,5].

The determinants of adolescent smoking are largely environmental. Parental and sibling smoking [6], exposure to secondhand smoke at home and in public places [4], and exposure to tobacco advertising, promotion and sponsorship [7,8] are consistently associated with initiation and use. The school setting holds a particular place: it can be a protective environment but also a place of exposure when adults, especially teachers, smoke there [9]. These factors nevertheless vary across cultural, regulatory and commercial contexts. They therefore need to be documented country by country to guide appropriate policies.

The Republic of the Congo has been a party to the WHO Framework Convention on Tobacco Control since 2007 [10]. It adopted a tobacco control law in 2012, supplemented in 2018 by decrees banning smoking in public places, all forms of tobacco advertising, promotion and sponsorship, and sales to minors [11,12]. In a country where young people make up approximately 60% of the population [13], data on adolescent tobacco use nonetheless remain scarce. Published studies have focused mainly on adult populations or specific groups in Brazzaville [14]. The third Global Youth Tobacco Survey (GYTS), conducted in 2019 on a nationally representative sample, provides an opportunity to analyse in depth the factors associated with tobacco use among Congolese middle-school students [15]. It also allows an indirect assessment of the implementation of this regulatory framework.

Beyond the national estimate, a simple tool could help school health teams identify students reporting exposures associated with current tobacco use. However, such a tool cannot predict future initiation, and its operational value must be assessed. The present analysis uses Congo GYTS microdata while distinguishing cross-sectional associations from identification performance within the existing data.

This study aimed to estimate the prevalence of current tobacco use among Congolese middle school students, to examine the factors associated with it, and to construct a simple identification score, assess its internal performance and compare it with that of statistical and machine learning models.

## Materials and methods

### Study design and population

This is a secondary analysis of data from the Global Youth Tobacco Survey (GYTS) conducted in the Republic of Congo in 2019. The GYTS is a cross-sectional, national, school-based survey that uses a standardised methodology developed by the WHO and the United States Centers for Disease Control and Prevention (CDC) [16]. The 2019 round was the third GYTS conducted in the Congo. It was implemented by the Ministry of Health and the Ministry of Education, with support from the WHO and the CDC. The anonymised public-use microdata were obtained from the WHO Noncommunicable Disease Microdata Repository [15] and accessed for research purposes on 26 March 2026.

Sampling followed a two-stage cluster design. Schools with probabilities proportional to enrolment were selected, and classes were randomly selected. All the students present in the selected classes were eligible to participate. The survey included 6,396 questionnaires (response rate 94.1%). The data file contains 34 strata and 70 primary sampling units (PSUs), as well as final weights (FinalWgt). Descriptive analyses cover students whose outcome status could be determined; models use complete observations for the variables included.

### Outcome and coding checks

The primary outcome was the use of at least one tobacco product in the past 30 days, based on CR7 (cigarettes, at least one day), CR10 (other smoked tobacco, “yes”) or CR14 (smokeless tobacco, “yes”). Shisha use (SR3), already included in other smoked tobacco in the GYTS questionnaire, was described separately without constituting a fourth product category. Electronic cigarettes were not included.

To reproduce the GYTS indicators, any student with at least one positive response was classified as a user; in the absence of a positive response, at least one valid negative response was sufficient to classify the student as a nonuser. Other observations were indeterminate and excluded. This rule yielded 6,297 usable observations; a strict rule, requiring three negative responses for nonusers, was examined in a sensitivity analysis. Reference results among 13–15-year-olds are reported separately.

### Variables

The adjusted model and the main score included eight factors defined before estimation: age, sex, exposure to smoke at home (CR19), exposure in enclosed public places (CR20), people smoking on school premises (CR22), teachers smoking at school (OR59), actual ownership of an item with a tobacco brand logo (CR37), and being offered a free tobacco product by someone working for a tobacco company (CR38). “Don’t know” responses concerning teachers formed a separate category. School grade was excluded from the main model to limit redundancy with age.

Willingness to use or wear an item with a tobacco brand logo (CR36) does not measure ownership. It was therefore reserved for a complementary analysis. Willingness to accept tobacco offered by a best friend (CR39), which may reflect behaviour already present, was not used in the main score. The other media and school variables were examined separately in crude analyses. Variable definitions and score coding are given in S1 Table.

### Statistical analysis

Proportions and their 95% confidence intervals (CIs) were weighted, with variances estimated by Taylor series linearisation over primary sampling units within strata. Crude and adjusted odds ratios (ORs) were derived from weighted logistic models. Standard errors were estimated via the complex-design sandwich matrix, with 36 degrees of freedom (70 primary sampling units minus 34 strata) and t-based intervals. The main multivariable model retained all prespecified factors without p value-based selection. The reporting follows the STROBE and TRIPOD guidelines [17–19].

Missing values of the predictors were described and compared between students with and without complete data. The main model was estimated on complete cases. A complementary analysis applied inverse probability weighting for being a complete case, modelled from smoking status and the available sociodemographic characteristics. This weighting assumes that data are missing at random, conditional on observed variables, and does not necessarily correct for unobserved biases.

To construct the score, each positive logistic coefficient was divided by a fixed unit of 0.30 and rounded to the nearest integer; negative coefficients were assigned zero points. The total score is the sum of the points. This scale was chosen to keep the tool short and easy to calculate. The classification thresholds were compared via the Youden index and the proportion of users identified. The principle of a point-based score is described by Sullivan et al. [20].

Discrimination was summarised by the weighted area under the ROC curve (AUROC). For internal validation, tenfold cross-validation was used, in which each primary sampling unit was assigned in its entirety to a single fold. Within each fold, the coefficients, and therefore the score points, were re-estimated on the training data only. Logistic regression, a CART tree and a random forest were compared using the same variables [21]. The 95% intervals for the AUROCs were obtained from 350 resamples of primary sampling units within strata. The out-of-fold calibration of the probabilities predicted by logistic regression was examined via the calibration slope, intercept and Brier score. The internal validation procedures followed those of Steyerberg et al. [22]. The Youden threshold was identified according to its original definition [23].

A complementary analysis reran the model including CR39; another replaced ownership (CR37) with willingness (CR36). A sensitivity analysis required all three product responses to be negative to define nonusers. Tables present unweighted counts and weighted measures, which are explicitly distinguished.

The analysis was performed in Python with NumPy, SciPy and scikit-learn. The estimation of regression coefficients and variance accounting for strata and primary sampling units was programmed explicitly and documented in the accompanying reproduction script. The pseudorandom seed for score development was set at 2019; resamplings for intervals and categories used distinct fixed seeds.

### Use of artificial intelligence tools

The authors used Claude (Anthropic), a large language model, to translate the manuscript from French into English, to draft and restructure parts of the Discussion, to adapt the manuscript to the journal’s format, to translate the labels of Fig 3 and to draw Figs 1 and 2 from values reported in the text and tables. The tool was not used for data collection or statistical analysis, and it did not generate any result. The authors checked all AI-assisted text, figures and references against the original French manuscript, the analysis outputs and the cited sources, and take full responsibility for the content of the article.

**Fig 1.**
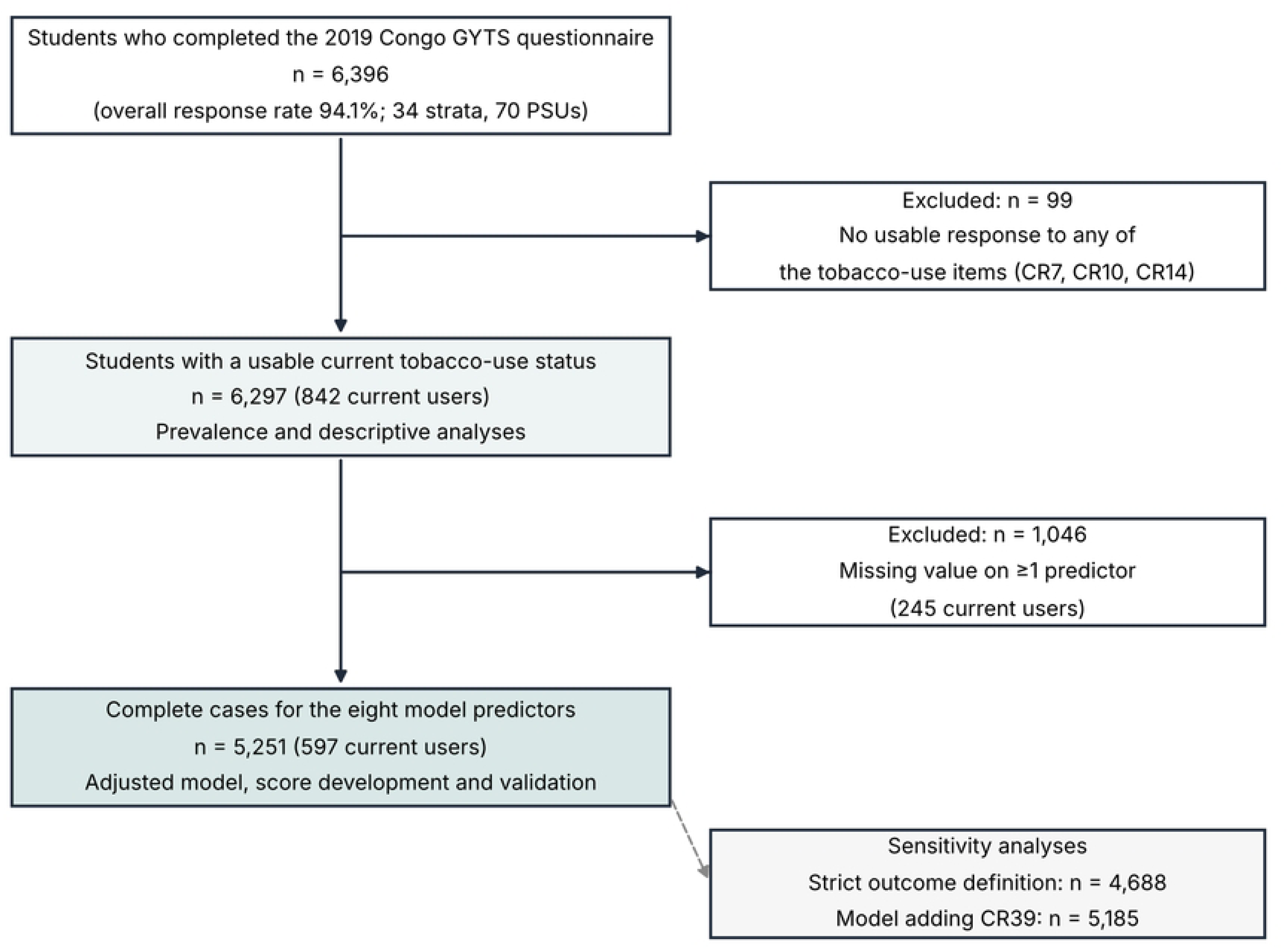
Flow diagram of study participants. GYTS: Global Youth Tobacco Survey; PSU: primary sampling unit. The diagram was drawn by the authors with AI assistance (Claude, Anthropic) from the participant counts reported in the text.

**Fig 2.**
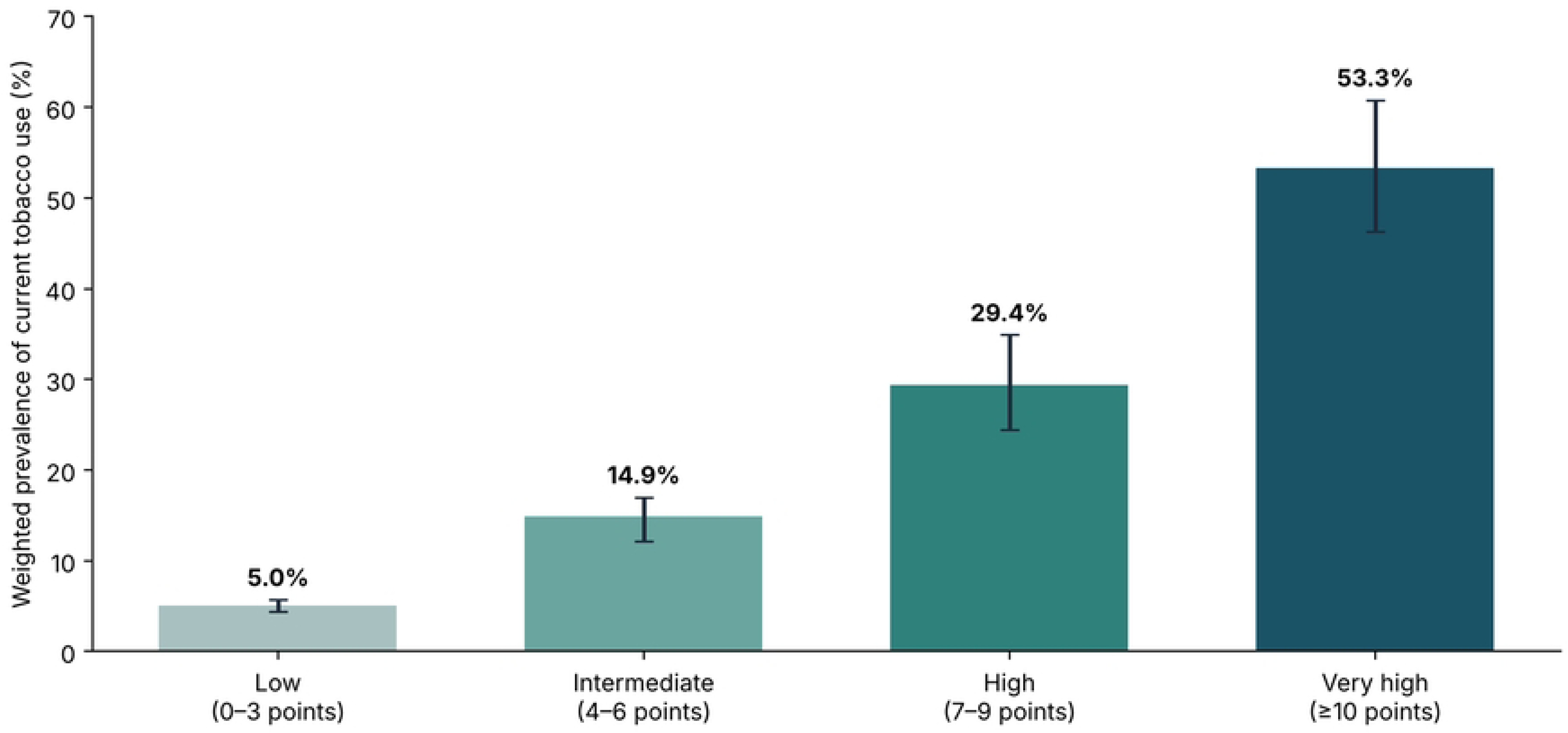
Weighted prevalence of current tobacco product use by score category. Error bars show 95% confidence intervals. The chart was redrawn with AI assistance (Claude, Anthropic) from the values in Table 5, which the authors verified.

**Fig 3.**
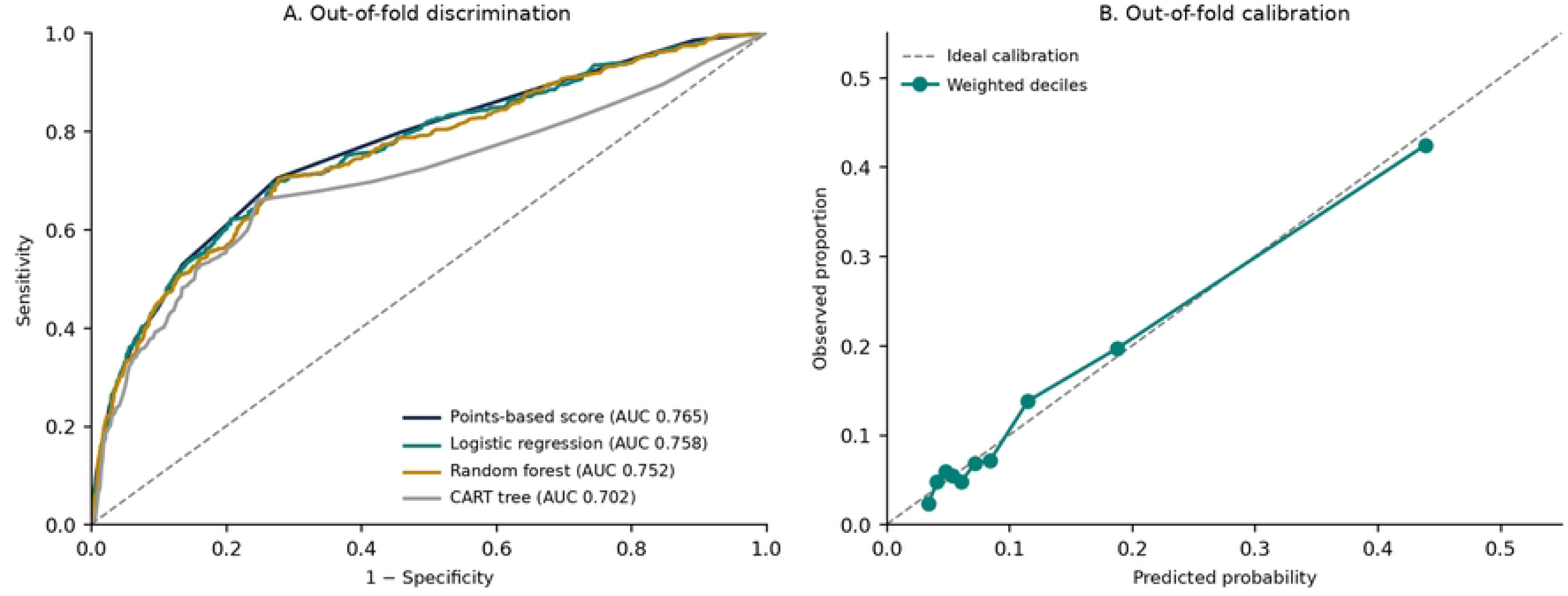
Out-of-fold discrimination and calibration of the models. (A) ROC curves of the point-based score, logistic regression, random forest and CART tree. (B) Observed versus predicted proportions by weighted decile for logistic regression. Models were re-estimated within cross-validation folds grouped by primary sampling unit. Axis and legend labels were translated from French with AI assistance (Claude, Anthropic); the plotted data were not modified.

### Ethics statement

The 2019 Congo GYTS was conducted by the Ministry of Health and the Ministry of Education of the Republic of Congo, with technical and financial support from WHO and the US CDC, according to the standard GYTS protocol, and was authorised by the national health and education authorities. Participation was voluntary and anonymous: the self-administered questionnaire did not collect names or any other information that could identify students. The present study is a secondary analysis of fully anonymised public-use data obtained from the WHO NCD Microdata Repository under its data access conditions; the authors had no access to information that could identify individual participants during or after data collection, and no additional ethical approval or consent was required for this analysis.

## Results

### Study population and prevalence of current use

Among the 6,396 participants, 6,297 had a usable response for the primary outcome, and 842 reported using at least one tobacco product (Fig 1). The weighted prevalence was 13.5% (95% CI: 11.1–16.3%). Among students aged 13–15 years, the prevalence was 14.1%, identical to the official GYTS estimate of 14.1% for this age group. The GYTS fact sheet used for international comparison covers 13–15-year-olds [24].

Cigarette use was 4.3%, the use of other smoked tobacco products was 7.1%, and smokeless tobacco use was 7.0%. Shisha use was reported by 3.3% of the students who answered this item. As product categories may overlap, their prevalence does not add up. Combining the three products of the standard indicator, 21.8% of current users reported at least two product categories, compared with 29.2% with the initial four-component breakdown, which potentially double-counted shisha.

Actual ownership of an item with a tobacco brand logo (CR37) was reported by 14.2% of the responding students; 25.9% reported being willing to wear or use such an item (CR36). These two measures are not interchangeable. Student characteristics and exposures are shown in Table 1.

**Table 1.** Descriptive characteristics of the population with a usable tobacco use status (N = 6,297).

| Characteristic | n/respondents | Weighted % | 95% CI |
| --- | --- | --- | --- |
| Age ≤12 | 1,669/6,228 | 22.7 | 19.3–26.6 |
| Age 13–14 | 2,631/6,228 | 42.7 | 39.6–45.9 |
| Age 15+ | 1,928/6,228 | 34.6 | 29.5–40.0 |
| Male | 3,140/6,153 | 50.9 | 47.9–53.8 |
| Female | 3,013/6,153 | 49.1 | 46.2–52.1 |
| Smoking at home | 600/6,196 | 10.1 | 8.1–12.6 |
| Smoking in enclosed public place | 1,121/6,162 | 18.4 | 15.7–21.3 |
| Smokers at school | 2,379/6,034 | 39.6 | 36.4–42.8 |
| Teachers smoking | 812/6,039 | 13.7 | 10.7–17.4 |
| Uncertain about teachers | 1,534/6,039 | 25.5 | 23.3–27.9 |
| Owns branded item | 760/5,932 | 13.3 | 10.8–16.2 |
| Willing to wear branded item | 1,482/6,102 | 25.6 | 22.7–28.7 |
| Offered tobacco by a company | 578/6,009 | 9.5 | 7.7–11.7 |
| Would probably accept tobacco from a friend | 346/6,098 | 5.9 | 4.9–7.1 |

Denominators vary according to valid responses for each question. Proportions and their CIs are weighted and account for stratification and cluster sampling.

### Factors associated in crude analyses

In the crude analysis, exposure to secondhand smoke, smoking at school and commercial promotion were associated with current use. The crude ORs were 6.29 for being offered free tobacco, 5.42 for exposure to smoke at home, 5.31 for teachers smoking at school and 4.53 for owning a branded item. These associations describe cross-sectional relationships and do not allow causal conclusions (Table 2).

**Table 2.** Crude associations with current tobacco product use.

| Variable/category vs reference | Users/n | Prev. (%) | Crude OR (95% CI) | p |
| --- | --- | --- | --- | --- |
| Age 13–14 years | 381/2,631 | 13.9 | 1.36 (0.98–1.90) | 0.066 |
| Age ≥15 years | 283/1,928 | 14.9 | 1.47 (1.03–2.12) | 0.037 |
| Boy | 489/3,140 | 15.1 | 1.48 (1.21–1.82) | < 0.001 |
| Smoke at home | 235/600 | 38.9 | 5.42 (3.73–7.89) | < 0.001 |
| Smoke in enclosed place | 332/1,121 | 30.0 | 4.04 (2.89–5.65) | < 0.001 |
| Smokers at school | 439/2,379 | 18.4 | 2.32 (1.80–3.00) | < 0.001 |
| Teachers smoking | 290/812 | 34.6 | 5.31 (3.86–7.30) | < 0.001 |
| Teachers unknown | 158/1,534 | 10.3 | 1.15 (0.89–1.47) | 0.278 |
| Owns branded item (CR37) | 267/760 | 33.6 | 4.53 (3.26–6.30) | < 0.001 |
| Free offer from a company | 231/578 | 41.2 | 6.29 (4.67–8.47) | < 0.001 |
| Would accept from friend: probably not (CR39) | 183/899 | 20.3 | 2.19 (1.74–2.76) | < 0.001 |
| Would accept from friend: yes (CR39) | 115/346 | 34.6 | 4.55 (3.22–6.43) | < 0.001 |
| Willing to wear branded item (CR36) | 415/1,482 | 26.9 | 4.10 (3.02–5.55) | < 0.001 |
| No anti-tobacco messages | 298/2,694 | 11.3 | 0.79 (0.63–1.00) | 0.048 |
| No teaching on tobacco | 190/1,718 | 11.0 | 0.62 (0.45–0.85) | 0.004 |
| Don't know about teaching | 220/2,098 | 10.7 | 0.61 (0.44–0.82) | 0.002 |
| Saw advertising | 244/1,255 | 19.6 | 2.53 (1.94–3.28) | < 0.001 |
| No visit to a point of sale | 305/1,850 | 16.1 | 1.99 (1.48–2.66) | < 0.001 |

Weighted ORs with design-based standard errors; p per category. Each factor was examined in a separate model; denominators differ according to missing values. The variables marked CR36/CR39 are not used in the main score.

### Adjusted associations and missing data

The main model included 5,251 students, of whom 597 were current users, compared with 6,297 observations for prevalence. Of the 1,046 students not included because of missing predictor values, 245 reported current use. The weighted prevalence was 11.5% among complete cases and 22.9% among those excluded. This difference suggests possible selection bias in complete-case models.

After adjustment, the strongest associations were with being offered free tobacco by a tobacco company (aOR = 3.05; 95% CI: 1.98–4.71), smoke exposure at home (2.71; 1.64–4.49), teacher smoking at school (2.60; 1.77–3.84), smoke exposure in enclosed public places (2.02; 1.40–2.93), and ownership of a branded item (1.73; 1.21–2.47). Male sex and age 15 years or older also remained statistically associated with current use (Table 3).

**Table 3.** Weighted logistic model and assigned points (n = 5,251; 597 users).

| Variable/category | Adjusted OR (95% CI) | p | Points |
| --- | --- | --- | --- |
| Age ≤12 years (ref.) | 1 (ref.) |  | 0 |
| Age 13–14 years | 1.22 (0.84–1.78) | 0.280 | 1 |
| Age ≥15 years | 1.69 (1.13–2.51) | 0.011 | 2 |
| Girl (ref.) | 1 (ref.) |  | 0 |
| Boy | 1.39 (1.04–1.87) | 0.030 | 1 |
| Smoke at home: no (ref.) | 1 (ref.) |  | 0 |
| Smoke at home: yes | 2.71 (1.64–4.49) | < 0.001 | 3 |
| Enclosed public place: no (ref.) | 1 (ref.) |  | 0 |
| Enclosed public place: yes | 2.02 (1.40–2.92) | < 0.001 | 2 |
| Smokers at school: no (ref.) | 1 (ref.) |  | 0 |
| Smokers at school: yes | 1.42 (1.10–1.82) | 0.008 | 1 |
| Teachers: never (ref.) | 1 (ref.) |  | 0 |
| Teachers: sometimes/daily | 2.60 (1.76–3.84) | < 0.001 | 3 |
| Teachers: don't know | 1.04 (0.78–1.38) | 0.799 | 0 |
| Branded item: no (ref.) | 1 (ref.) |  | 0 |
| Branded item: owns (CR37) | 1.73 (1.21–2.47) | 0.004 | 2 |
| Free offer: no (ref.) | 1 (ref.) |  | 0 |
| Free offer: yes | 3.05 (1.98–4.71) | < 0.001 | 4 |

Points derived from positive β coefficients/0.30, rounded. The “don’t know” coefficient for teachers receives zero points. The scale ranges from 0 to 18 points; all the predictors are retained in the model.

### Score construction and performance

The coefficients of the full model were converted into a score ranging from 0 to 18 points. Free tobacco offered by a tobacco company contributed four points, smoke at home and teachers smoking three points each; the other items received between zero and two points (Table 3). The apparent AUROC was 0.771 (95% resampling CI: 0.743–0.793). In cross-validation grouped by primary sampling unit, the AUROC of the score was 0.765 (95% CI: 0.736–0.788).

The threshold for maximising the Youden index in the development population was ≥4 points, with a weighted sensitivity of 70.9%, specificity of 71.8%, positive predictive value of 24.6% and negative predictive value of 95.0%. At this threshold, 33.2% of the students were classified as positive (Table 4). The ≥3-point threshold identified more users (sensitivity 82.2%) but had a specificity of 50.8%. These threshold values were calculated on the basis of the development data and must be evaluated prospectively.

**Table 4.**
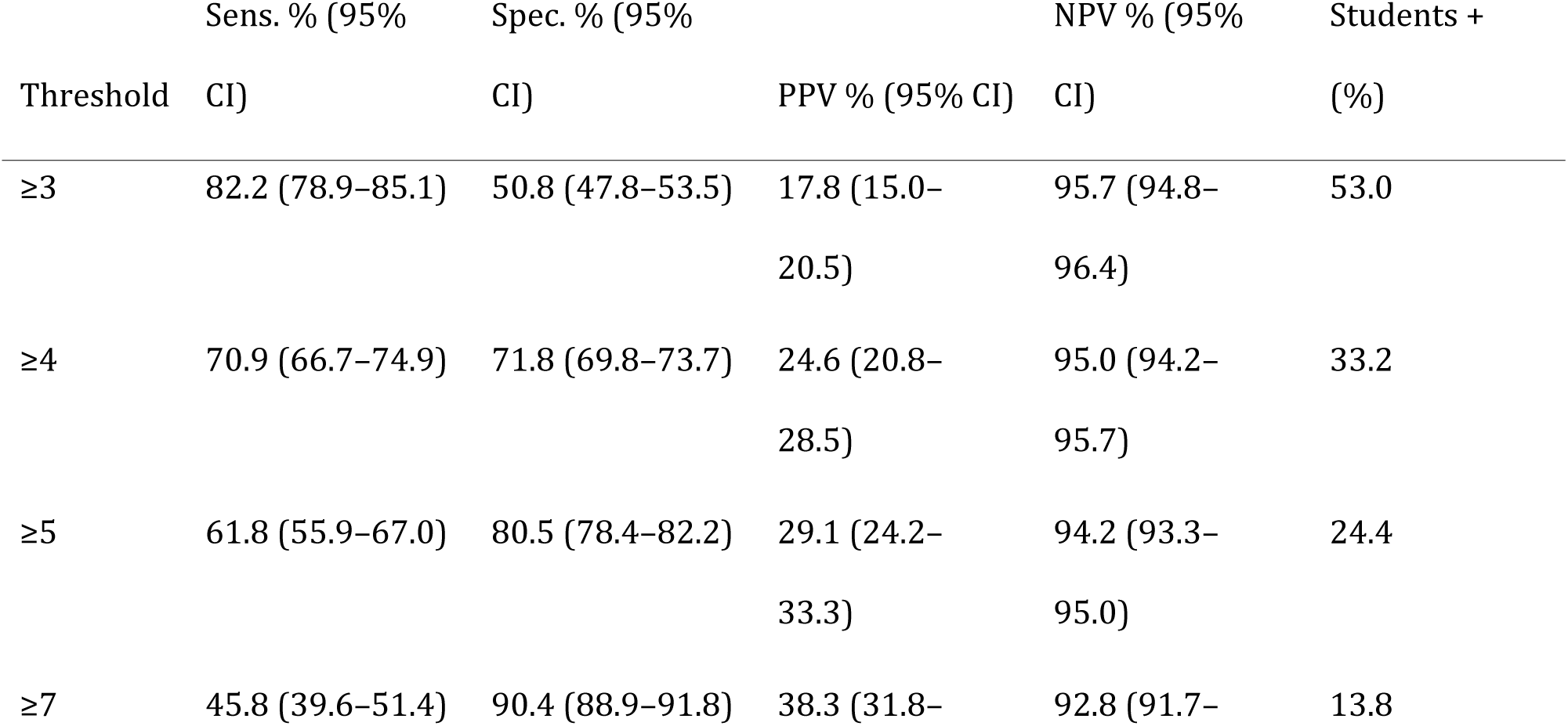
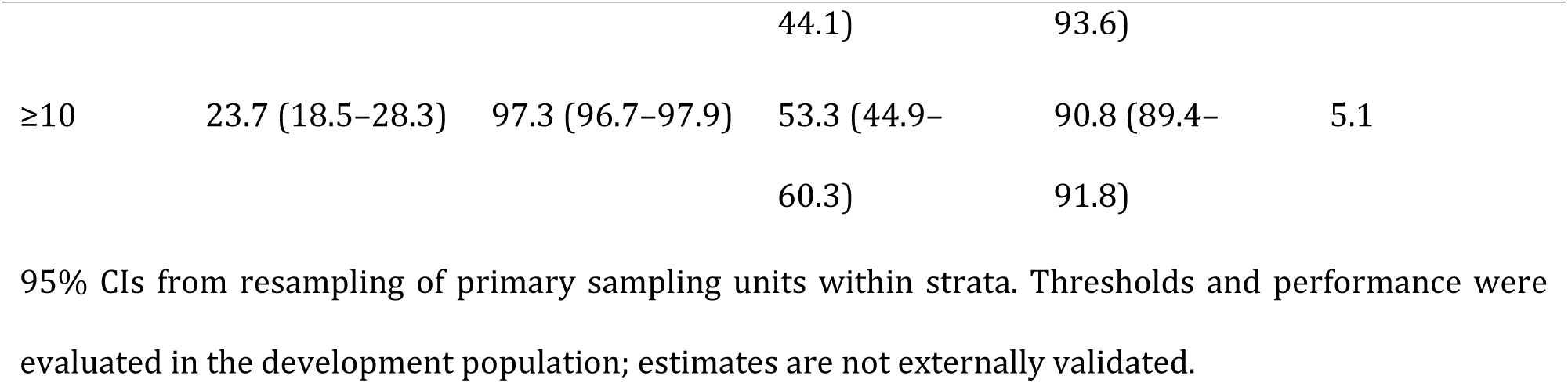
Weighted classification performance by threshold.

### Stratification and comparison of methods

The weighted prevalence increased from 5.0% for a score of 0–3 points to 53.3% for a score of at least 10 points. The latter category represented 5.1% of the students in the model (Table 5 and Fig 2). These groups reflect observed consumption, not future risk.

**Table 5.**
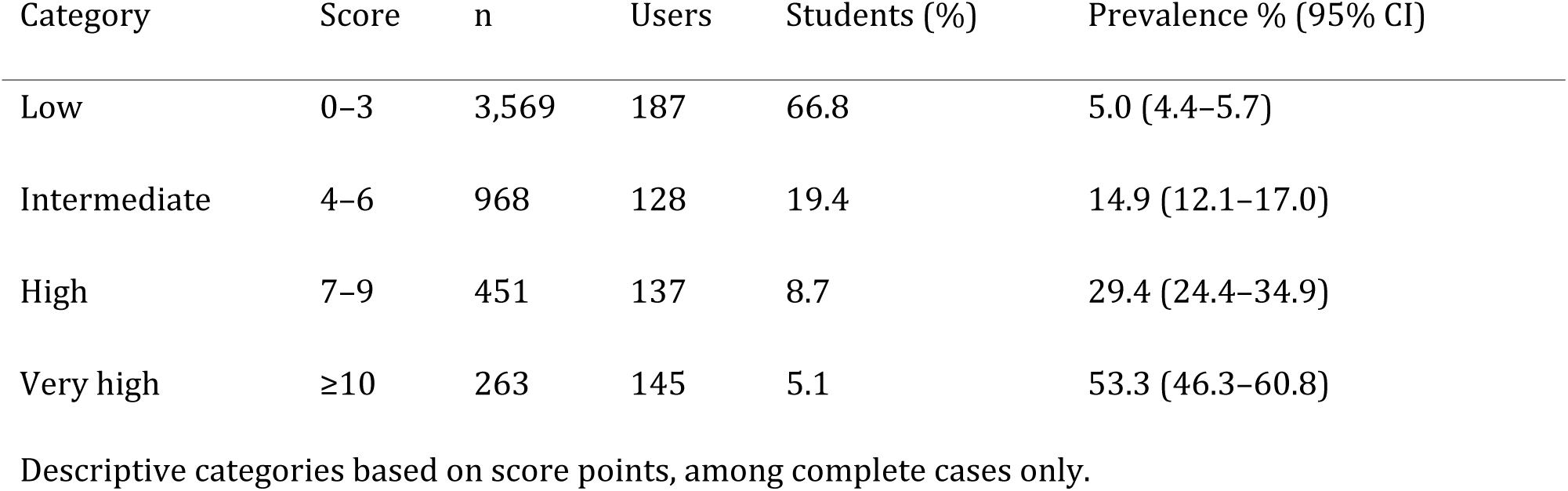
Current tobacco use by score category.

In cross-validation grouped by primary sampling unit, the weighted AUROCs were 0.765 for the score, 0.758 for logistic regression, 0.752 for the random forest and 0.702 for the CART tree. The random forest offered no clear gain over the score. For logistic regression, the out-of-fold calibration intercept was −0.09, the slope was 0.95, and the Brier score was 0.088 (Table 6 and Fig 3).

**Table 6.** Comparison of methods by tenfold grouped cross-validation.

| METHOD | WEIGHTED AUROC | 95% CI |
| --- | --- | --- |
| Points-based score | 0.765 | 0.736–0.788 |
| Logistic regression | 0.758 | 0.731–0.783 |
| Random forest (250 trees) | 0.752 | 0.725–0.776 |
| CART tree (depth 4) | 0.702 | 0.668–0.730 |

In each fold, the parameters were re-estimated on the training primary sampling units. Intervals were obtained by resampling primary sampling units within strata for out-of-fold predictions.

### Sensitivity analyses

When nonusers were required to answer negatively to all three components, 4,688 complete-case students were retained; the weighted prevalence was 12.9%, and the apparent AUROC of the model was 0.790. Inverse probability weighting for completeness yielded ORs of the same order as the main model for the main exposures (Table 7). These analyses do not rule out bias related to missing values.

**Table 7.** Robustness of adjusted ORs under different analytical assumptions.

| Variable | Complete cases | Completeness weighting | Strict outcome |
| --- | --- | --- | --- |
| Smoke at home | 2.71 | 2.66 | 2.99 |
| Smoke in public place | 2.02 | 2.05 | 2.42 |
| Smokers at school | 1.42 | 1.43 | 1.35 |
| Teachers smoking | 2.60 | 2.66 | 3.43 |
| Owns branded item | 1.73 | 1.74 | 2.02 |
| Free offer | 3.05 | 3.15 | 4.05 |

Strict outcome: 4,688 complete cases; negative status only if all three responses were negative Completeness weighting: inverse probability weighting estimated from smoking status and sociodemographic characteristics. These analyses are exploratory.

Adding willingness to accept tobacco from a friend (CR39) increased the apparent AUROC to 0.784 among 5,185 complete cases. Replacing actual ownership (CR37) with willingness to wear a branded item (CR36) yielded an apparent AUROC of 0.791. These analyses were not used to construct the main score.

## Discussion

According to the analysis of the 2019 GYTS, 13.5% of the Congolese middle school students reported using tobacco in the past 30 days. In the 13–15-year-old subgroup, the prevalence was 14.1%, which is consistent with the WHO national fact sheet [24]. This value is lower than the 19.1% reported by James et al. in 22 African countries [4] and the 18.6% reported by Itanyi et al. in Nigeria [5]. However, the years, populations and products studied differ. Surveillance should not be limited to cigarettes: smokeless tobacco and other smoked products also contribute to adolescent consumption.

Reported exposure to smoke at home (aOR = 2.71) and in enclosed public places (aOR = 2.02) was associated with use. James et al. reported associations in the same direction in Africa, with values of 1.98 and 1.37, respectively [4]. Leonardi-Bee et al. associated family smoking with youth initiation [6]. The questionnaire specifies neither who smoked nor the cumulative duration of exposure. History-taking should specify the products used, frequency, age at initiation, smoking habits of those around the adolescent and quit attempts, distinguishing passive exposure from active use.

The school environment deserves attention. Seeing teachers smoke was associated with current use (aOR = 2.60). Al-Binali et al. reported a weaker association in Iraq (aOR = 1.48) [9]. Being offered free tobacco by a tobacco company was our strongest association (aOR = 3.05), a value also observed for industry promotion by James et al. [4]. In Nigeria, Chido-Amajuoyi et al. associated cumulative exposure to advertising with current cigarette use (aOR = 1.73) [8]. The measures differ, but they call for enforcing bans on promotion and smoking at school.

From a clinical perspective, these associations do not demonstrate any individual harm. The GYTS collected no physical examination, auscultation, or respiratory or cardiovascular diagnosis. Neither cough, wheezing, dyspnoea nor clinical dependence can be estimated. For a student reporting tobacco use, a confidential consultation could look for these manifestations, assess the frequency of use and offer age-appropriate advice. A further clinical examination would be indicated for adolescents presenting with respiratory complaints or seeking help to quit, ensuring confidentiality and the absence of stigmatisation.

No measurements of cotinine, exhaled carbon monoxide or lung function were available. Self-reports expose the study to misclassification. Valladolid-López et al. reported, among 1,257 Mexican adolescents, a sensitivity of 93.2% and a specificity of 81.7% for GYTS self-reports compared with urinary cotinine [25]. Ab Manan et al. reported a sensitivity of 67.6% and a specificity of 99.3% in Malaysia [26]. A consented substudy could compare questionnaire responses with those of salivary or urinary cotinine. This biomarker reflects nicotine exposure and depends on secondhand smoke and the products used. Targeted spirometry could be offered in the presence of persistent symptoms without serving as biological proof of use.

Our 0–18-point score achieved a weighted AUROC of 0.765 in cross-validation grouped by primary sampling unit. At the four-point threshold, its positive predictive value was 24.6%. The score recognises profiles associated with reported use; it predicts neither future initiation nor disease. However, its advantage over a direct question remains unknown. Before any use in schools, external validation and an acceptability study are needed. This validation should include urban and rural schools from several regions.

### Strengths and limitations

This study has several strengths. It is based on a nationally representative sample of more than 6,000 middle-school students, collected via the standardised GYTS methodology, with a high response rate (94.1%). All estimates account for the complex sampling design (weights, strata and primary sampling units), allowing valid inference at the national level. The outcome follows the standard WHO definition, which facilitates international comparisons; the prevalence obtained among 13–15-year-olds is identical to the official estimate. Model factors were defined a priori, without p value-based selection, and variables likely to reflect use itself were excluded. Finally, internal validation of the score, by cross-validation grouped by primary sampling unit with re-estimation of points in each fold, limits optimism in the reported performance.

These results must nevertheless be interpreted in light of several limitations. First, the cross-sectional design does not establish the temporality of associations: some exposures, such as owning a branded item or being offered free tobacco, may be a consequence of use rather than its cause. Second, use and exposures were self-reported and not biologically verified; underreporting due to social desirability, particularly in a school setting, is possible. Third, models were estimated on complete cases; excluded students reported current use twice as often (22.9% vs 11.5%), and inverse probability weighting for completeness did not correct for biases related to unobserved factors. Fourth, the GYTS covers only students enrolled in school and present on the survey day; out-of-school or frequently absent adolescents, who are probably more exposed, are not represented. Fifth, the data date from 2019 and reflect neither possible changes since the COVID-19 pandemic nor e-cigarette use, which is excluded from the standard definition. Finally, the score has not been externally validated: its thresholds were determined on the development data, and it identifies students who already report tobacco use and does not predict future initiation. Its value compared with a direct question about consumption remains to be demonstrated.

## Conclusions

Current tobacco use remains common among Congolese middle-school students. Exposure to smoke and commercial promotion practices are associated with this consumption. The proposed identification score is a first methodological step, whose relevance in schools must still be established through external validation and a field study.

## Data Availability

No data was generated by this study. The following existing data sources were used: the 2019 Congo Global Youth Tobacco Survey (GYTS) dataset, obtained from the WHO NCD Microdata Repository and available via who.int.

https://www.google.com/url?sa=i&source=web&rct=j&url=CAESdQHrOzAVaVF98HgIXF9Et1zUN27xNzB3vCQ5Zqh1ueENZBgrWiCZ6MQck8Q1a0ezh5JFNR36ox8Sz-xLsyzTSqLFUmtj_jK1ezrtE59LyFUgC75N0OTKqvslg6Yu9ZL3vPvaL7kbTGjFeAJ1hljxACs4hA2hpQ&uoh=2&ved=2ahUKEwiT1N_unr6XAxWVV0EAHX8KAzYQy_kOegYIAAgSEAE&opi=89978449&cd&psig=AOvVaw2GlmGPdJn8Wtl7QHWKETR6&ust=1792229919149000

## Acknowledgments

This study uses data from the Global Youth Tobacco Survey (GYTS), which is supported by the World Health Organization and the U.S. Centers for Disease Control and Prevention. The authors thank the Congolese Ministry of Health and Ministry of Education for conducting the survey.

## Supporting information

**S1 Table. Variables in the main model and score coding.** Definitions of the eight predictors, their categories and the points assigned in the score.

**S1 Checklist. STROBE checklist for cross-sectional studies.**

